# Household food insecurity, living conditions, and individual sense of security among Burkina Faso refugees in Ghana: a cross-sectional survey using in-person data collection

**DOI:** 10.1101/2023.07.31.23293476

**Authors:** Abdul-Wahab Inusah, Ken Brackstone, Michael Head, Tahiru Issahaku Ahmed, David Tetteh Nartey, Shamsu-Deen Ziblim

**Affiliations:** JSI Research & Training Institute, INC. Takoradi, Ghana; Clinical Informatics Research Unit, Faculty of Medicine, University of Southampton, UK; School of Medicine, University for Development Studies, Northern Region, Ghana; F.N. Binka School of Public Health, University of Health and Allied Sciences, Hohoe, Volta Region, Ghana; Bawku West District Assembly, Upper-East region, Bolgatanga, Ghana; Directorate of Academic Planning & Quality Assurance (DAPQA) University for Development Studies, Tamale, Ghana

**Keywords:** Household food insecurity, Burkina Faso refugees, Food security, Living conditions

## Abstract

Food insecurity and adequate nutrition is a major global challenge, especially in vulnerable groups such as refugee communities. In West Africa, due to conflict and instability in their home country, thousands of Burkina Faso refugees have crossed the border into northern Ghana. We conducted a cross-sectional study aimed at assessing household food insecurity, living conditions, and sense of security among Burkina Faso refugees currently residing in the upper east region of Ghana. Study data was collected from 19-29 October 2022 from 498 refugee households. Results revealed that almost 100% of households experienced food insecurity, with 80.3% experiencing severe food insecurity. Refugees from rural areas were more food secure compared to urban refugees. Refugees residing in host communities experienced lesser food insecurity than those in designated refugee camps. Further, refugees who were dissatisfied with the size of their accommodation were more likely to experience food insecurity. More than half of the refugees felt safe and welcomed by the host communities. The study underscores the critical need for targeted intervention to avert hunger and malnutrition among vulnerable groups such as children, pregnant women, and the aged in the refugee population. The study also emphasizes the importance of community support systems and social amenities for refugees’ well-being. Long-term measures should include improving logistics for food distribution, providing suitable accommodation, and strengthening healthcare services. Follow-up research, such as repeated community surveys, can track this evolving situation to continuously inform decision-making for refugee support.

## Introduction

Food insecurity leads to malnutrition, which negatively impacts upon all individuals, but can have a devastating effect on more vulnerable groups such as young children and pregnant women (1,2). The UN estimates that 735 million people, around 9% of the world’s population, are experiencing hunger and food insecurity, with increases in the prevalence of undernourishment observed in sub-Saharan Africa(3). The Global Nutrition Report highlights that no African country is on track to meet any diet-related non-communicable disease targets. This makes it challenging, if not impossible, to achieving the SDG target to eradicate hunger by 20230(4).

Globally, approximately 26 million people are currently refugees with close to 80% of this population experiencing food insecurity regardless of location (1). Refugees often flee from armed conflict. Violence can directly lead to food insecurity and malnutrition by obliterating food systems, reducing farming labour, and destroying transport systems, thus leading to a reduction in community resilience (5–7).

Burkina Faso is in West Africa, with an estimated population of 22.1m. It is a landlocked nation bordering several other countries, including Ghana to the south. Since 2015, Burkina Faso has witnessed several waves of armed conflicts emanating from non-state armed groups whose aims are to instrumentalize communal tensions and exploit the vulnerability of an already fragile population. This has resulted in a displacement surge among its citizens [1]. Reports from UNICEF indicate that Burkina Faso recorded 488 armed attacks with 144 people killed in the first quarter of 2022 (8). As a result of this conflict, the number of internally displaced persons (IDP) has risen to over 1.8 million, with 61% of the group being children (9). The ongoing instability and violence means that there are declining crop yields, with a continued rise in prices of goods and services (8). As a result, around 13 million people are faced with food insecurity, and one-third are malnourished children aged under 5 years old (10).

The insecurity situation in the country has resulted in 179 healthcare centres being closed in the hardest hit regions, with 353 healthcare centres operating at suboptimal levels. These closures have denied over 2 million citizens access to health care (8). A significant proportion of population now have little access to basic social amenities such as water, food, shelter, education, and healthcare, with many people considering that their best option is to flee to neighboring countries. As of February 2023, Ghana hosted 13,500 refugees, of which 4000 of these were Burkina Faso citizens seeking asylum (11). These numbers continued to increase across 2022 and 2023, particularly within the Upper East region of Ghana, which directly borders Burkina Faso (8). The care of these refugees is the sole responsibility of the Ghanaian local and national authorities and is supported by international humanitarian agencies such as United Nations High Commissioner for Refugees (UNHCR). Their arrival has not only resulted in increased pressure on Ghanaian and international agencies to provide humanitarian needs for these refugees, but also places excess pressure on social amenities in their host communities.

This study was conducted to assess the health, social, and economic needs of these refugees in the Upper East region of Ghana, with a specific focus on food insecurity and the living conditions among this population. We aimed to explore and understand predictors of food insecurity, and to describe some of the key characteristics of this vulnerable group.

## Methods

### Study design and settings

A community-based cross-sectional study was implemented in Ghana using in-person data collection from household heads of identified refugees (supplementary S1). Data was collected in the rural districts of Binduri District, Bawku Municipal, and Bawku West, situated within the Upper East region of Ghana (Figure 1), where the majority of displaced people are located. The region contains 15 administrative districts with a population of 1,301226 (12), and a total landmass of 8,842sq/km constituting 2.7% of the country’s land size (13). The regional capital is Bolgatanga, approximately 60-100km west of the study sites. Upper East region shares international borders in the north with Burkina Faso, and in the east with Togo (13). The location of the region exposes it to a lot of cross-border activities with neighbouring migrants and traders from Burkina Faso and Mali, Togo and Niger using it as transmit point into Ghana.

**Fig 1.**
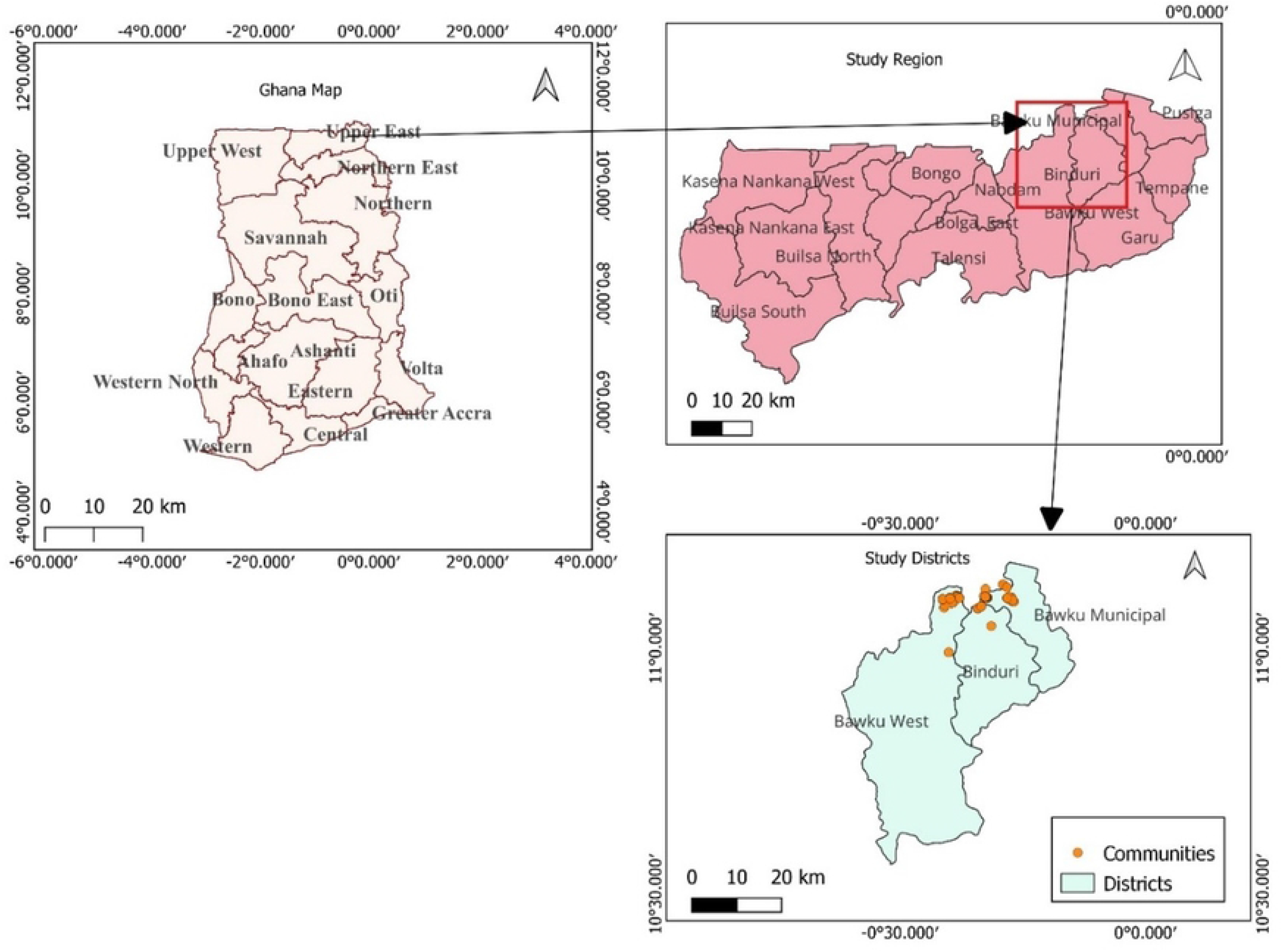
Map showing the regions of Ghana, Upper-East region, and the municipals with refugee communities. Image created in QGis Version 3.28.1 by author Abdul-Wahab Inusah.

### Study population and sample size determination

All consenting adult heads of refugee households in the host communities were eligible participants. Start and end dates of data collection was 11-21 October 2022. Sample size was calculated using a single population proportion formula with an assumption of 50% population proportion (p = 0.5) due to the unknown population size. With a 95% confidence level, a 5% margin of error, and a normal population distribution Z = 1.96, we adjusted for a 5% non-response rate. This provided an estimated sample size of 400.

### Sampling technique and sampling procedures

Using the list provided by the National Disaster Management Organization department of all three administrative districts, the number of participants studied in each district was proportional to the total number of registered refugees in the district. The number of registered refugees for each district was 1072 in Binduri District (64%), 495 in Bawku Municipal (29%) and 112 in Bawku West district (7%), which totalled 1679 individuals. Since the calculated sample size for the study was 400, the number of participants for each district was 256 for Binduri district, 116 for Bawku Municipal, and 28 for Bawku West district. A purposive sampling technique was used to select only communities hosting Burkina Faso refugees. With the help of the community refugee focal persons (persons appointed by the respective local assemblies) who documented all refugees in the communities, we identified and contacted all registered refugee households using the refugee register for each community. In each household, the household head was our respondent. A household was revisited in the absence of the household head. If that individual was still not available upon the second visit, the survey was carried out with an individual nominated from the rest of the household members.

### Study variables

#### Household food insecurity assessment

We used the USAID household food insecurity assessment score (HFIAS) module (14), which was used to assess household food insecurity over the previous 30 days. HFIAS consist of 9 questions grouped into 3 domains, including: (1) anxiety and uncertainty, (2) insufficient quality, and (3) insufficient food intake and physical consequences. Participants responded “yes” or “no” to each of the nine questions. Those who answered “yes” were further asked about the frequency of the occurrence of the condition and the responses were” rarely”, “sometimes”, or “often”. We summed the responses to the nine questions to get the HFIAS index. The minimum score was 0 if the participant responded “no” to all the nine conditions and the maximum of 27 if all the nine conditions were selected “yes”. A higher HFIAS score indicates greater food household insecurity and a lower score indicates greater food security (14). Households were finally categorized into secure, mildly food insecure, moderately food insecure, and severely food insecure consistent with the HFIAS score (14).

#### Living condition variables

Participants were asked about their current accommodation, and living conditions including whether they knew locals prior to arriving in the community, the size of their accommodation facility, and presence of any social support for their household and sources of these social support systems.

#### Sense of security

To measure the safety and sense of security among the study participants, 4 questions were asked using the 5-point Likert scale (e.g., “Generally, I feel satisfied with my current living conditions”; 1 = *strongly disagree*; 5 = *strongly agree*). We averaged the items to form a single index (*M* = 3.46, *SD* = 0.99, α = 0.68).

#### Sociodemographic variables

These included: sex (male, female), age (continuous), marital status (not married, married, separated/divorced, widowed), religious affiliation of the household head (Christian, Muslim, Traditional, other), household size (number of people sharing a common shelter), number of children under five in the household, and number of household members pregnant.

#### Data collection procedure and quality control

Some of the questions were adapted from an existing questionnaire used in 2022 with Ukrainian refugees and internally-displaced persons by the same research team (15,16). The survey questions were uploaded to an Android smartphone App (ODK). The tool was pretested in Nabdam, a neighbouring district among 20 refugees. These test responses were not included in the final analysis. All the necessary adjustments, including skip logic, and age groupings were made before the main data collection begins. Ten community health workers participated in a two-day data collection training workshop, covering issues such as data collection, good research practice and the need for explaining the study and obtaining informed consent. As we anticipated low literacy rates, survey questions were verbally read out and the data collectors recorded their responses directly onto the ODK app. The survey was written in English. A local dialect, Moshie, was used for verbal translations and context.

### Data analysis techniques

Stata version 15 was utilized in carrying out descriptive and inferential statistics for the data. Descriptive statistics including frequencies, percentages, mean and standard deviation. A reliability test was conducted for the 9 HFIAS and the 4 sense of security variables and a Cronbach alpha value of 0.84 and 0.68 respectively were obtained. To assess determinants of household food insecurity, a logistic regression analysis was conducted, and p-values of < .05 were considered statistically significant.

### Ethical consideration

Ethical approval (UDS/RB/112/22) was obtained from the University for Development Sciences Research and Ethics Review (Supplementary, S2). Separately, approval and community entry was sought from the 3 district assemblies to carry out the study within their populations.

Informed written consent was obtained from all individuals before they participated in the study.

## Results

In total, 498 participants completed the survey (Table 1), with a majority being female (65.3%). The average age was 37.2 years (*SD* = 16.2, *Range* 18-98). Nearly 86.0% had no formal education, 93.0% were affiliated to Islamic religion, and 90.0% indicated that their home community was a rural area (Table 1). The mean household size was 5.97 people, with a mean of 1.42 children aged under 5 years per household. Finally, 5.5% of the female group indicated that they were pregnant.

**Table 1.** Sociodemographic Characteristics of the Burkina Faso Refugees in Ghana (*n* = 498)

| Characteristics | Frequency(n) | Percentage (%) |
| --- | --- | --- |
| **Sex** |  |  |
| Female | 325 | 65.3 |
| Male | 173 | 34.7 |
| **Age (Years)** |  |  |
| 18-30yrs | 227 | 45.6 |
| 31-40yrs | 115 | 23.1 |
| 41-50yrs | 45 | 9.0 |
| 50-60yrs | 57 | 11.4 |
| 61yrs+ | 54 | 10.8 |
| **Marital status** |  |  |
| Never married | 35 | 7.0 |
| Divorced/widowed | 70 | 14.1 |
| Married | 393 | 78.9 |
| **Religious affiliation** |  |  |
| Christian | 29 | 5.8 |
| Traditional | 7 | 1.4 |
| Islam | 462 | 92.8 |
| **Educational level** |  |  |
| No formal education | 426 | 85.5 |
| Basic secondary school | 63 | 12.7 |
| Complete secondary school | 9 | 1.8 |
| **Description of participants home community** |  |  |
| Countryside/rural | 448 | 90.0 |
| Urban/city | 50 | 10.0 |
| **When did you arrive at your current location?** |  |  |
| 1-7 days ago | 9 | 1.8 |
| 2 weeks-1month ago | 60 | 12.1 |
| More than 1month ago | 429 | 86.1 |

### Prevalence of household food insecurity among the study households

Of the 498 households served, only 4 (0.8%) of households were defined as food secure, with 494 (99.2%) of households defined as experiencing mild to severe food insecurity (Table 2, Figure 2). Across the previous 4 weeks, 35% of households were often worried that their household did not have enough food, and approximately 38% of households were not able to eat the kind of food the household preferred. Results also show that 27.5% of households have a member/s who had to go through a whole day and night without eating anything because there was not enough food available.

**Fig 2.**
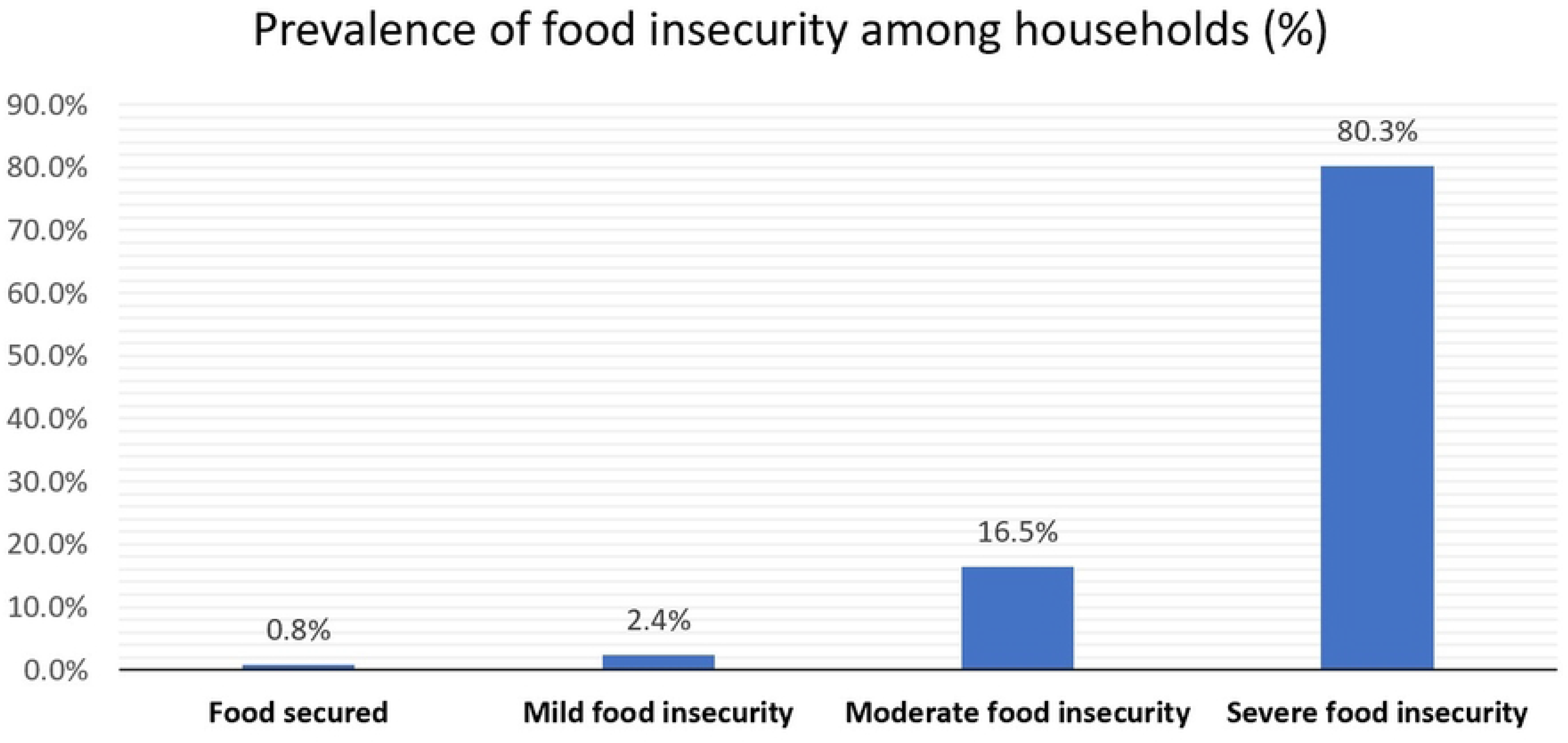
Overall prevalence of household food insecurity among the study population

**Table 2.** Household Food Insecurity Access Scale (HFIAS) components among households in the past 4 weeks.

| HHFS Items | Never<br>Experienced | Rarely<br>Experienced | Sometimes<br>Experienced | Often<br>Experienced |
| --- | --- | --- | --- | --- |
| Q1 In the past four weeks, did you worry that your household would not have enough food? | 36(7.2) | 66(13.3) | 224(45.0) | 172(34.5) |
| Q2 In the past four weeks, were you or any household member not able to eat the kinds of foods you preferred because of a lack of resources? | 36(7.2) | 53(10.6) | 221(44.4) | 188(37.8) |
| Q3 In the past four weeks, did you or any household member have to eat a limited variety of foods due to a lack of resources? | 19(3.8) | 63(12.7) | 259(52) | 157(31.5) |
| Q4 In the past four weeks, did you or any household member have to eat some foods that you did not want to eat because of a lack of resources | 18(3.6) | 76(15.3) | 232(46.6) | 172(34.5) |
| Q5 In the past four weeks, did you or any household member have to eat a smaller meal than you felt you needed because there was not enough food? | 15(3.0) | 49(9.8) | 265(53.2) | 169(33.9) |
| Q6 In the past four weeks, did you or any other household member have to eat fewer meals in a day because there was not enough food? | 56(11.2) | 47(9.4) | 226(45.4) | 169(33.9) |
| Q7 In the past four weeks, was there ever no food to eat of any kind in your household because of a lack of resources to get food? | 55(11.0) | 62(12.4) | 206(41.4) | 175(35.1) |
| Q8 In the past four weeks, did you or any household member go to sleep at night hungry because there was not enough food? | 42(8.4) | 129(25.9) | 165(33.1) | 162(32.5) |
| Q9 In the past four weeks, did you or any household member go a whole day and night without eating anything because there was not enough food? | 92(18.5) | 99(19.9) | 170(34.1) | 137(27.5) |

### Determinants of household food insecurity

From a simple linear logistic regression model, the sociodemographic variables were not significant (Table 3). We therefore adjusted for only three significant variables, such as refugee residents before migrating to their current location, refugees’ current location in Ghana, and the size of the accommodation facilities in the final multivariate logistic regression model. From the model, rural refugee were negatively associated with food insecurity. That is, refugees from rural areas were more food secure than those from cities or urban areas, B = -4.25 (CI: -5.79 – -2.71), *p* < .001. Refugees currently staying with someone they knew, such as a relative or a friend in the host community, reported lower food insecurity compared to those living in housing specifically set up to house refugees, B = -1.56 (CI: -2.74 – -0.39,) *p* = .009. Finally, lower satisfaction with accommodation space was marginally associated with increased food insecurity compared to those happy with the size of their accommodation space, B = 2.96 (CI: -0.06 – 2.47), *p* = .060. Results are displaced in Table 3.

**Table 3.** Multivariate logistic regression model of predictors of food insecurity among Burkina Faso refugees in Ghana (n = 489)

| Variable | B | t | P-value | [95% CI] |
| --- | --- | --- | --- | --- |
| Resident before migrating to Ghana |  |  |  |  |

| City/Town | Ref |  |  |  |
| --- | --- | --- | --- | --- |
| Countryside/rural | -4.25 | -5.4 | 0.001* | -5.79 – -2.71 |

**Current residence (Ghana)**
| In housing specifically set up to house refugees<br>or internally displaced people | Ref |  |  |  |
| --- | --- | --- | --- | --- |
| With a local person or people who have offered<br>me accommodation | 2.34 | 1.5 | 0.10 | -0.35 – 2.43 |
| With somebody I knew before, such as relatives<br>or friends | -1.56 | -2.6 | 0.009* | -2.74 – -0.39 |
| With somebody with whom I was put in touch,<br>for example, friends of friends | -0.53 | -0.4 | 0.66 | -2.92 – 1.86 |

**How would you describe the size of your accommodation?**
| Happy with the space I have currently | Ref |  |  |  |
| --- | --- | --- | --- | --- |
| Least happy with the space I have | 1.2 | 1.8 | 0.06 | -0.062 – 2.47 |

### Living conditions of the refugees

By accommodation status, 54.0% were living in housing specifically set up to house refugees, with 22.7% living with somebody they knew before migrating to Ghana, such as relatives or friends (Table 4). A further 19.5% were living in accommodation provided by a member of the host community who was not previously known to the refugee participant. Most females (69.1%) were living in housing specifically set up to house refugees (**Error! Reference source not found.**). By living conditions, 84.1% of participants were least happy about their current living conditions, with only 2.4% reporting being happy. In terms of support system, only 120 (24.1%) participants had received any welfare or housing payment to support their living standards (Table 4). Of the 120 participants who received financial support, most had received support from more than one source, with 94.2% receiving welfare payments from the Ghanaian Government, and 85.0% from international agencies such as UNICEF and UNHCR as their source of welfare payments (Figure 3). Overall, 76.0% of respondents indicated they did not have enough financial and other resources to meet their household basic needs.

**Fig 3.**
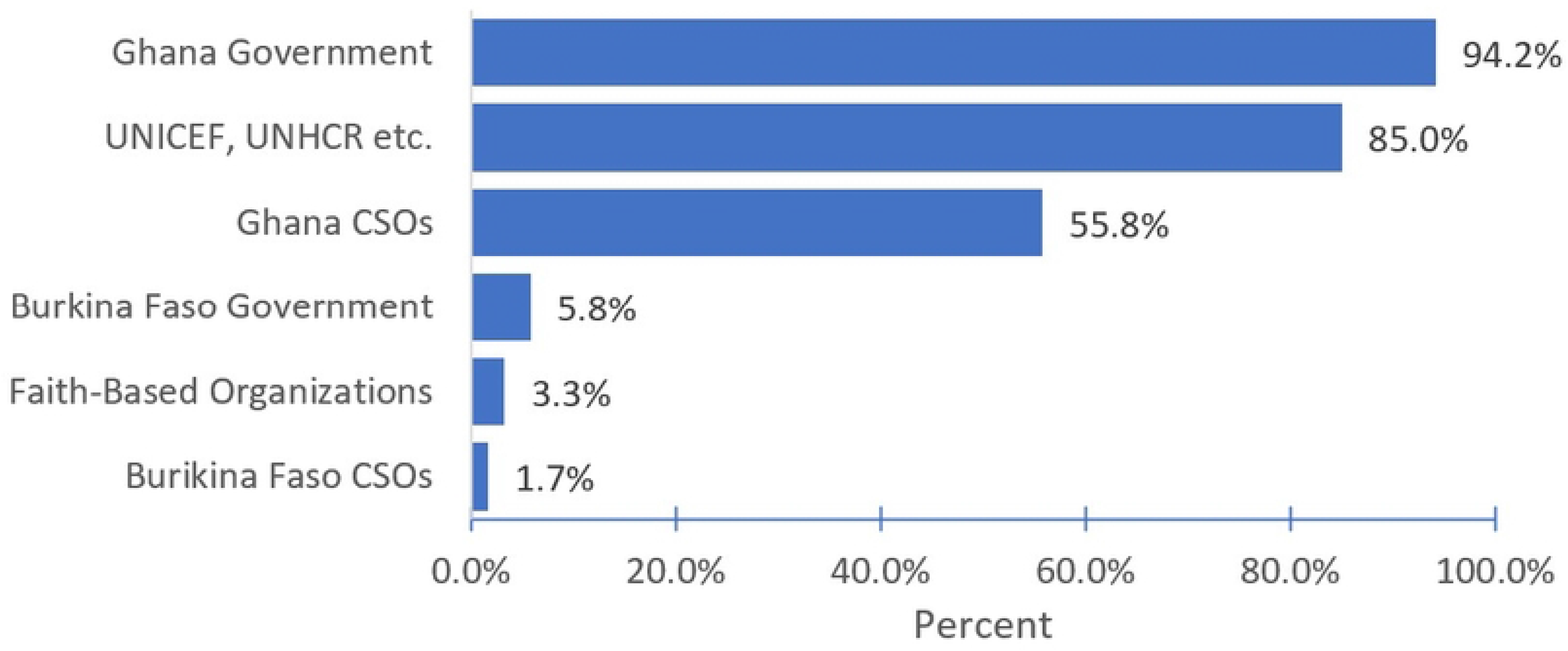
Main Sources of Welfare or payment support among Burkina Faso Asylum refugees in Ghana (*n* = 120)

**Table 4.**
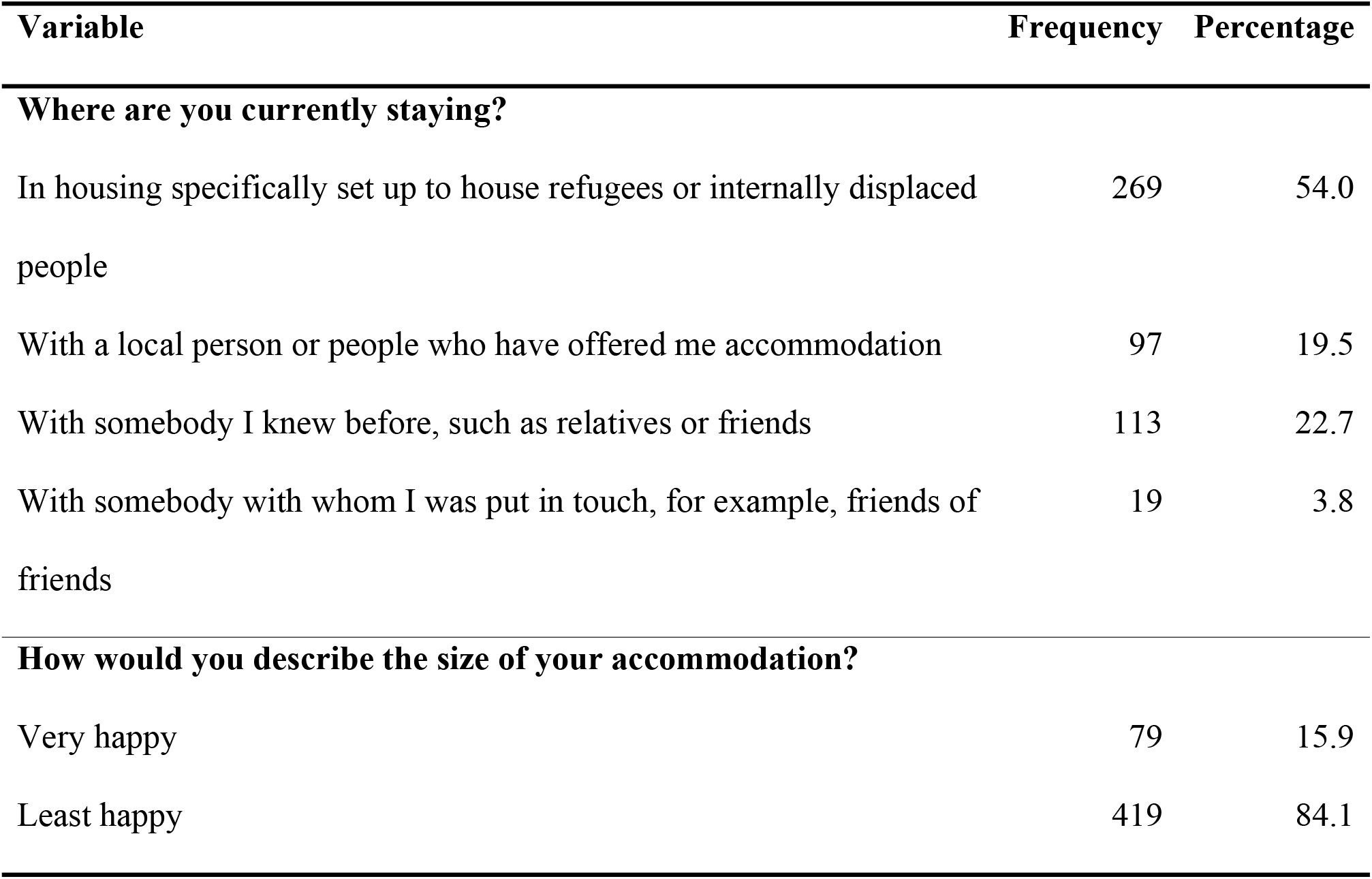
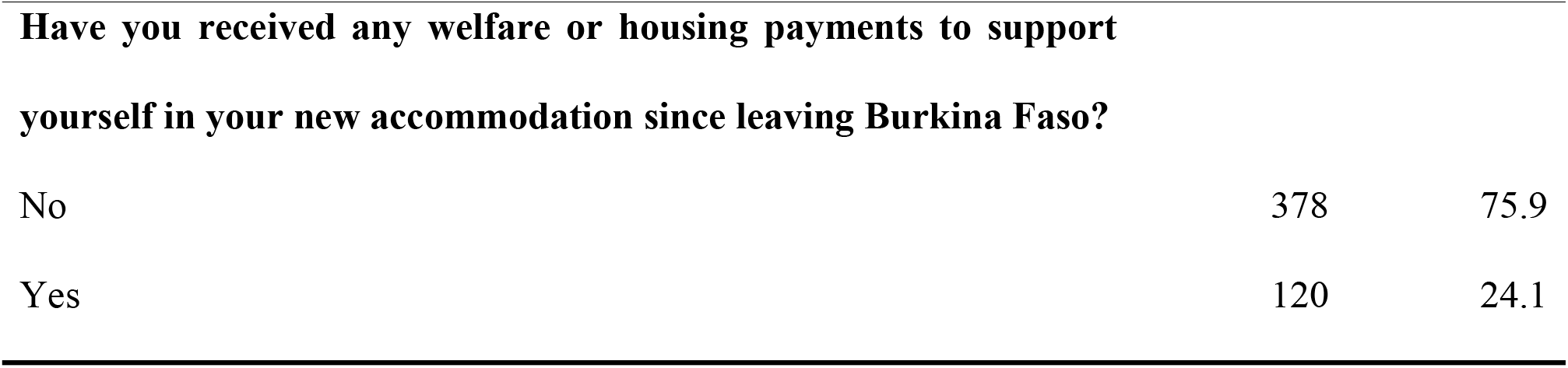
Living Conditions of the Burkina Faso refugees in Ghana (n = 498)

### Self-reported sense of security in the current living conditions among Burkina Faso Refugees

Regarding participants’ sense of security, 71.9% of respondents agreed to some extent that they feel satisfied with their current living conditions. However, 28.5% of participants disagreed to some extent that they feel satisfied with their current living conditions. With regards to community safety, 72.5% agreed to some extent that they felt safe in their new location, with 16.0% expressing some form of disagreement. By community response, 73.5% agreed to some extent that they felt welcome by the local community, whereas around 17.0% disagreed to some extent. Finally, participants 23.3% indicated that they had perceived some level of hostility or anger from others in their local community (Table 5).

**Table 5.** Sense of security in the current living conditions of the study participants.

| Variables | Responses (%) |  |  |  |  | Mean |
| --- | --- | --- | --- | --- | --- | --- |
|  | Strongly disagree | Somewhat disagree | Uncertain | Somewhat agree | Strongly agree |  |
| Q13 Generally, I feel satisfied with my current living conditions | 72 (24.5) | 20 (4.0) | 48 (9.6) | 129 (25.9) | 229 (46.0) | 3.8 |
| Q14 I feel safe in the place where I am currently | 54 (10.8) | 26 (5.2) | 57 (11.4) | 95 (19.1) | 266 (53.4) | 4.0 |
| Q15 I have been made to feel welcome by the local community | 49 (9.8) | 20 (4.0) | 63 (12.7) | 76 (15.3) | 290 (58.2) | 4.1 |
| Q16 My family and I have experienced hostility or anger from other people in the local community | 338 (67.9) | 22 (4.4) | 22 (4.4) | 63 (12.7) | 53 (10.6) | 1.9 |

## Discussion

Displaced persons are often disadvantaged with access to adequate food and suitable living conditions. Among Burkina Faso refugees currently living in the Northern part of Ghana, our study found that close to 100% experienced household food insecurity, with 80.3% of households experiencing severe insecurity. Self-reported satisfaction of living conditions was variable across the study population, with most participants reporting how they felt safe and welcomed by their new local community.

The level of food insecurity was higher than the national prevalence of food insecurity in both Burkina Faso (41%) and the host country Ghana (11.7%) (17,18). Although, a recent study found some level of food insecurity reported by Ghanaian participants from a rural community in the Northern Region of Ghana(19). Among other refugee populations, food insecurity is typically very high. For example, several studies have reported greater than 80% insecurity for refugees in the USA, with 42% of children experiencing hunger (20). Similar results were found among Syrian refugees in Canada (21), and Afghan refugees in Iran (22). Lower insecurity was reported among Iraqi refugees in Lebanon (44.4%) and Scotland (56%) (23). Our findings add to the evidence base around risks of hunger and malnutrition among these vulnerable populations.

There may be a need for targeted interventions to reduce the negative impact on the lives of under-5s, pregnant women, and older people, as these populations are disproportionately affected by food insecurity and conflict (21,24,25) For those who remained in Burkina Faso, there is a longer-term trend of difficulties with food security as a result of climate change, the impact of COVID-19, increases in food prices, and the continued rise in armed conflicts (24,26,27).

Around half of the participants in this study resided in refugee camps set up by UNHCR and the local authorities, and these people reported less satisfaction and greater insecurity with the quality of their accommodation compared to participants who were staying with community members who they knew. This may highlight the increased vulnerability of forced migrants who have little knowledge of their new community, with issues such as familiarity and language being factors that may make it more difficult to access food, water, and healthcare.

Most participants (80%) reported some level of unhappiness around the size of their accommodation. A qualitative study conducted in Southern Australia among refugees and asylum seekers exploring the relationship between housing and health revealed that a significant number of participants were worried about the size of the accommodation which was reported to have a negative impact on their mental health (28). There are many factors that affect wellbeing of refugee groups, and this provides an opportunity for stakeholders to conduct site-specific needs assessments to determine optimum and realistic living conditions alongside the level of satisfaction among refugees.

Refugees are entitled to receive welfare support from local authorities, although this is often difficult due to individuals’ lack of familiarity with bureaucratic and regulatory barriers. From our study, only 24.1% of refugees received welfare support with the major source of accessible support coming from Government of Ghana and the United Nations agency, such as UNICEF and UNHCR. This finding indicates the key role played by host-country in supporting the mandated UN systems to take care of displaced persons during emergencies.

Around half of the participants indicated that they felt safe and welcomed by the host communities. Refugees’ sense of safety and security in the host community is a complex issue that varies across different contexts. Studies have shown that host communities’ attitudes toward refugees can have a significant impact on their sense of safety and security (29). Acceptance and assistance from the host community are crucial in ensuring support for refugee populations (30). However, the perception of safety among refugees is subjective and individual, with many refugees not considering themselves to have found complete safety and protection in their host countries (31).

Our analyses indicate that refugees who migrated from rural areas had higher levels of food security compared with those from urban areas. The majority of participants indicated they were originally from rural areas. Thus, it is possible that rural-dwellers can more easily assimilate themselves with community members for support compared to those from cities who may have a problem adjusting to rural life. This is consistent with an earlier study conducted in Australia, which revealed that refugees from rural communities found it easy assimilating with rural host communities compared to urban refugees (32). There can also be a negative impact upon food security within the host communities themselves, as an influx of refugees can lead to uneven distribution of limited food supplies (29). In this study, refugees who are presently residing with an acquaintance within the host community, such as a relative or a friend, encountered fewer complexities concerning food insecurity in comparison to individuals residing in housing exclusively designated for refugees or internally displaced persons. These findings are in line with an earlier study reporting the importance of social support and community networking in improving the well-being of refugees (33,34). The result equally confirms the trade and inter-personal relationships that exists between the Northern part of Ghana and southern Burkina Faso (35). These relationships are established by historical, cultural, and economic elements, with migration and familial connections present between the populations of both nations (36).

The number of the displacement of Burkina Faso populations are on a much smaller scale than the situation with Ukraine, and perhaps this is one of the reasons for the lower profile of this crisis in West Africa. However, the needs and conditions of the populations described here appear to be worse than those experienced by Ukraine refugees who are typically wealthier and moving to higher-income settings. When comparing Burkina Faso refugees in Ghana with the population remaining within Afghanistan, the prevalence of food insecurity is similarly high.

Malnutrition is one of the key concerns in Afghanistan, and this has to be a real concern for those addressing the refugee crisis in northern Ghana.

## Strengths and limitations

The strength of this study lies in the fact that this was a community-based cross-sectional study, with high-level refugees’ participation. This study fills a knowledge gap around assessment of food insecurity, safety, and wellbeing among refugees within the sub-region.

However, the study contains limitations. The tool that was used to measure household food insecurity provides a direct measure of household ability to afford food within the last 30 days and does not take into account other factors, including seasonality of food production and availability, discrimination in food allocation, and food preference. These aspects must be explored further to understand how they influence household food insecurity. There is little data available around the demographics of this refugee population, and thus we cannot be sure how representative our study populations are. This is also a one-off survey at one point in time – the refugee situation evolves quickly, and repeated similar measurements would provide an up to date assessment of health and social needs.

## Conclusion

The current study found a high prevalence of household food insecurity among the Burkina Faso refugee population residing in Ghana. Refugees staying with populations known to themselves reported higher food security. Participants who were unhappy with their accommodation size were associated with a higher risk of household food insecurity. A majority of the participants felt safe and welcomed by the host communities. The short-term measures should include improved logistics for regular food distribution by governments, UNHCR, and UNICEF. Measures should be taken to improve the living conditions through the provision of acceptable accommodation facilities. Health services should be strengthened, with proactive review of potential emerging cases of malnutrition. Further research can include repeating this survey to ensure up-to-date findings are available for decision-makers.

## Data Availability

The dataset is publicly available at https://doi.org/10.6084/m9.figshare.23782893

## Acknowledgements

The study team would like to acknowledge and thank the participants and data collectors for their help and support with this research.

## Supporting information

S1. Survey administered to the Burkina Faso refugee participants S2. Confirmation of ethics approval

